# Emergence and Circulation of a Recombinant Enterovirus D68 Identified by Genomic Surveillance, The Johns Hopkins Health System, Maryland, 2025

**DOI:** 10.1101/2025.11.22.25340766

**Authors:** Amary Fall, C. Paul Morris, Omar Elgazayerly, Andy Wu, Omar Abdullah, Julie M. Norton, Andrew Pekosz, Eili Klein, Heba H. Mostafa

## Abstract

**Background:** Enterovirus D68 (EV-D68) is an important respiratory pathogen occasionally linked to acute flaccid myelitis. While recombination drives enterovirus evolution, recombinant EV-D68 strains have been rarely documented.

**Methods:** As part of 2025 genomic surveillance in Maryland, 115 EV-D68 genomes were sequenced using an amplicon-based approach. Consensus genomes were aligned with global references and analyzed with IQ-TREE3 and SimPlot to assess phylogeny and recombination.

**Results:** Complete genomes were obtained from 78% (90/115) of specimens, all belonging to subclade A2. Five genomes formed a distinct cluster with discordant phylogenies across genomic regions: P1 grouped with A2, whereas P2–P3 clustered with B3. SimPlot and BootScan analyses identified a recombination breakpoint near the 2A/2B junction (∼nt 3,700), consistent with an A2(P1)/B3(P2–P3) recombinant. BAM alignment review excluded co-infection.

**Conclusions:** We report a novel EV-D68 A2/B3 recombinant circulating locally in 2025, highlighting the need for continued whole-genome surveillance.

## 1. Introduction

Enterovirus D68 (EV-D68), a member of the *Picornaviridae* family, has re-emerged as a significant human pathogen over the past decade. In contrast to most enteroviruses, EV-D68 primarily targets the respiratory tract, leading to disease manifestations from mild upper-respiratory infection to severe respiratory distress requiring hospitalization (1-3). Of public-health concern is its established association with acute flaccid myelitis (AFM), a neurological disease characterized by acute limb weakness and paralysis, predominantly in children (3-6).

The evolution of EV-D68 is marked by the global co-circulation of multiple genetic clades. Following the major 2014 outbreak driven by subclade B1, subsequent epidemic waves have shown diversification within clades A and B, with subclades B2 and B3 predominating in recent years (1, 5, 7-10). This diversification is fueled by two principal evolutionary forces: the accumulation of point mutations and genetic recombination (11-15). Recombination is a well-recognized mechanism of enterovirus evolution, allowing exchange of genomic fragments between related viruses during co-infection of a host cell (16-18). Such events can give rise to novel lineages or subclades with altered pathogenicity and fitness or antigenic characteristics that could pose challenges for both surveillance and outbreak control (19-22).

Although recombination reportedly contributes to enterovirus evolution, only rare sporadic recombinant events have been reported for EV-D68 (23-27). Identification and characterization of these recombinant genomes are critical for understanding the full scope of the virus’s evolutionary dynamics and adaptive potential.

During our ongoing genomic surveillance of enteroviruses in Maryland in 2025, we identified a cluster of EV-D68 sequences that could not be definitively assigned to established subclades based on whole-genome analysis, suggesting a recombinant origin. Here, we describe the genomic detection and confirmation of a novel recombinant EV-D68 strain.

## 2. Methods

### 2.1. Sample Collection, Nucleic Acid Extraction and EV-D68 screening

Clinical specimens were obtained through the Johns Hopkins Health System (JHHS) enterovirus genomic surveillance research in 2025. Remnant respiratory samples that tested positive for rhinovirus/enterovirus (RV/EV) using the ePlex respiratory pathogen panels were collected for further analysis (approved IRB protocol IRB00221396). Viral RNA extraction was performed with the Chemagic Viral DNA/RNA 300 Kit. Samples were screened for EV-D68 by real-time reverse transcription polymerase chain reaction as described previously (7, 28, 29).

### 2.2. Whole Genome Sequencing and sequences analysis

Whole-genome sequencing was performed as previously described (1, 10). Amplicons from two overlapping PCR reactions yielding fragments of approximately 4,380 bp and 3,200 bp respectively were pooled and barcoded using the Native Barcoding Genomic DNA Kit (EXP-NBD196; Oxford Nanopore Technologies) according to the manufacturer’s instructions. Sequencing was conducted on a PromethION 2 Solo (P2) (Oxford Nanopore Technologies) device equipped with R10.4.1 flow cells. Raw FASTQ files for each sample were generated directly by the P2 and processed using a custom in-house pipeline. Primer sequences were trimmed with cutadapt (version 3.5)(30), and the closest reference genome was identified by BLASTn (31) against a curated Enterovirus database. Reads were aligned to the reference genome using minimap2 (version 2.30)(32), then sorted and indexed with Samtools (version 1.10)(33). Consensus genomes were generated and polished with medaka (Version 2.1.1)(34) . Variants were filtered with ARTIC tools using custom versions of artic_vcf_filter, artic_make_depth_mask, and artic_mask (35). Final consensus sequences were generated using bcftools (36).

### 2.3. Phylogenetic analysis

Complete genomes were aligned with reference genomes from GenBank using Mafft (version 7.450). Maximum likelihood trees for complete genomes were carried out using IQ-TREE3 (version 3.0.0), with 1000 bootstrap replicates. The ModelFinder, implemented in IQ-TREE3, was used to select the best-fitted nucleotide substitution model.

### 2.4. Recombination analysis

Recombination detection was performed using multiple complementary approaches to ensure reliability. Whole-genome alignments of representative Enterovirus D68 strains were generated with MAFFT (v7). Phylogenetic incongruence between the P1, P2 and P3 regions was further assessed using maximum-likelihood trees constructed in IQ-TREE3 with 1,000 bootstrap replicates. To visualize recombination breakpoints and confirm parental relationships, SimPlot (v3.5.1) was used to perform similarity and Bootscan analyses using a 200-bp sliding window and 20-bp step size. In addition, BAM alignment files were inspected in Integrative Genomics Viewer (IGV) (V2.19.6) to assess possible co-infections or mixed subclade signals within the samples analyzed. Sequencing of two samples was repeated to confirm the reproducibility of our results.

## 3. Results

A total of 977 samples positive for RV/EV collected between June and October 2025 were screened for EV-D68. Among these, 115 tested positive for EV-D68 with a cycle threshold (Ct) value ≤35 and were selected for sequencing. Complete genomes were successfully recovered from 78% (90/115) of samples, while partial genomes were obtained from an additional 6% (7/115) of specimens.

Phylogenetic analysis of complete genomes confirmed the exclusive circulation of the A2 subclade in Maryland during 2025, with all sequences clustering within this lineage **(Figure 1)**. Notably, five genomes formed a distinct, monophyletic cluster that branched near A2 references. This emerging cluster was named A2-Re and exhibited shorter branch lengths relative to contemporary A2 sequences, a signature of reduced genetic divergence that often may point to a recent recombination event. These five sequences were also detected in different geographic areas.

**Figure 1.**
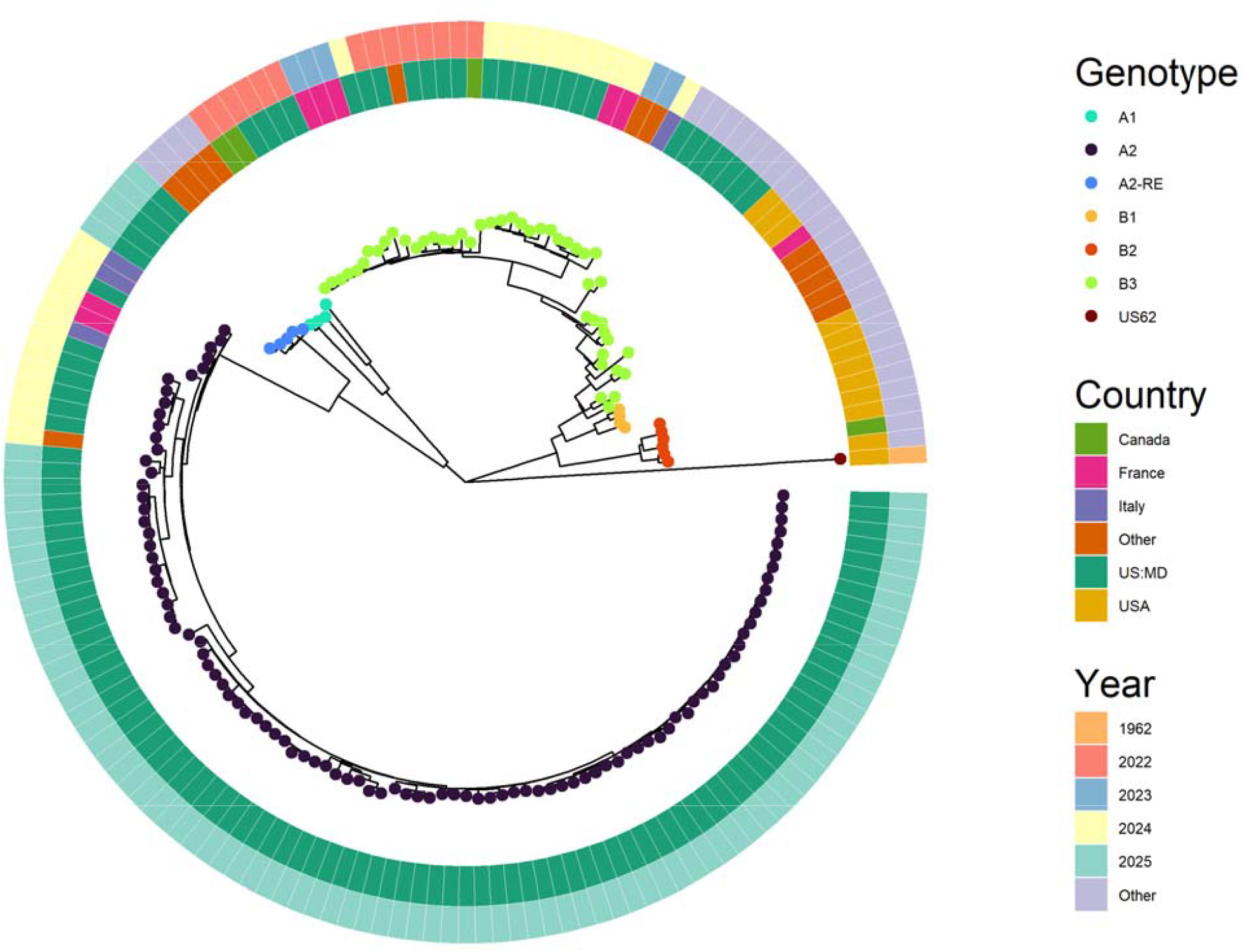
Phylogenetic relationships of EV-D68 strains identified from the Johns Hopkins Health system in 2025. Complete genome sequences from other countries were included. The phylogenetic tree was constructed using complete genomes, employing the maximum likelihood method in IQ-TREE2 with 1,000 bootstrap replicates and rooted by the Fermon strain.

To investigate this possibility, we constructed subgenomic trees for the P1 (structural), P2, and P3 (non-structural) regions. We supplemented each dataset with closely related sequences identified via BLAST. These analyses revealed clear topological incongruence. In the P1 tree, the cluster grouped tightly with 2024 and 2025 A2 viruses, consistent with the whole-genome phylogeny (Figure 2). In contrast, the P2 and P3 trees placed these same genomes within the B3 subclade, forming a well-supported cluster with viruses that circulated in Canada and the United States in 2022 (Figure 2). Within the P2 tree, the cluster exhibited longer branch lengths than typical B3 sequences, suggesting this region may be a chimera derived partly from an A2 lineage or has accumulated unique mutations. This shift in phylogenetic placement between the structural and non-structural regions indicates a recombination event, in which the P1 region was derived from an A2 parental strain, whereas part of the P2 region and the entire P3 region originated from a B3 lineage.

**Figure 2.**
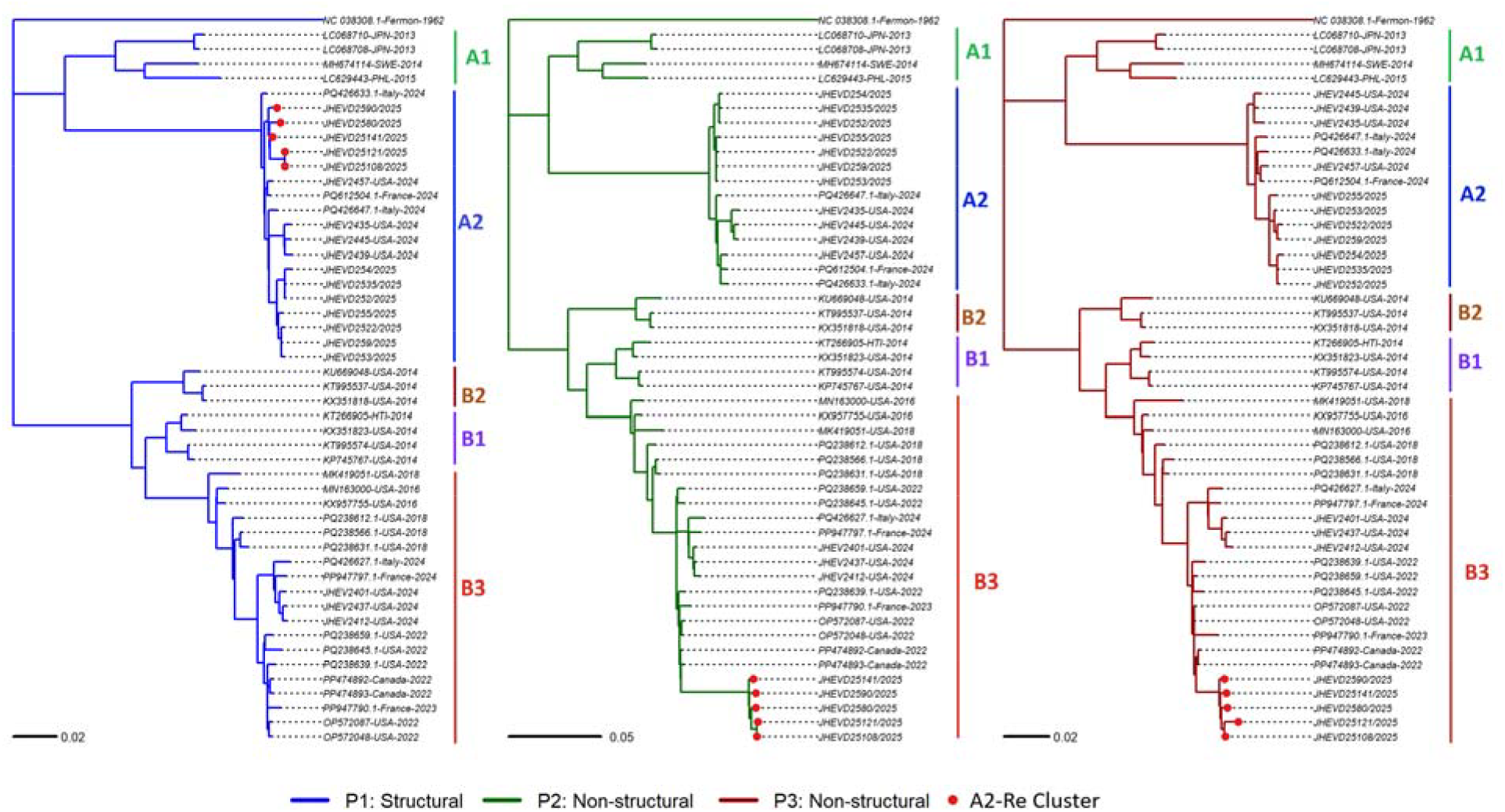
Phylogenetic analysis of the P1, P2, and P3 coding regions of EV-D68 strains from this study and globally available reference sequences. Maximum-likelihood phylogenetic trees were inferred using IQ-TREE3 from alignments of approximately 2,586 nt (P1, blue), 1,728 nt (P2, green), and 2,256 nt (P3, red).

To further confirm the recombination event and precisely locate the breakpoint, SimPlot analysis revealed a recombinant genome structure characterized by alternating regions of high sequence similarity to A2 and B3 reference strains **(Figure 3A)**. The plot displayed a sharp transition in similarity profiles near nucleotide position 3,700, corresponding to the 2A/2B junction

**Figure 3.**
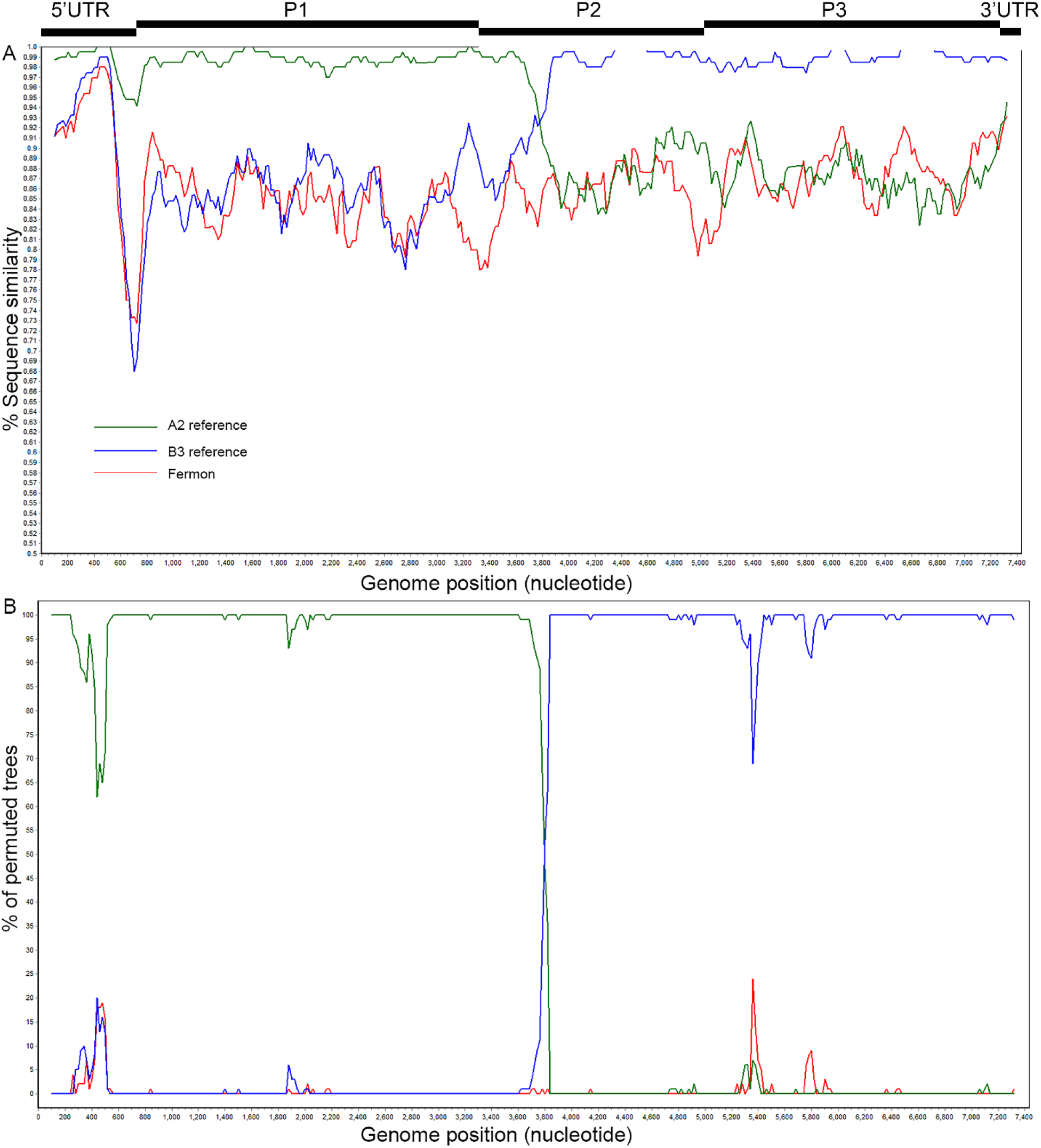
Recombination analysis of EV-D68 A2/B3 recombinant genomes using SimPlot and BootScan. Similarity (A) and bootstrap support (B) plots were generated with SimPlot v3.5.1 using a 200-bp sliding window and a 20-bp step size. The recombinant Maryland sequences (A2-Re_2025) show high similarity to A2 strains from Maryland (2025) in the P1 (structural) region and to B3 strains from Canada and Maryland (2022) in the P2–P3 (non-structural) regions, with a clear recombination breakpoint near nucleotide position 3,700.

This pattern was corroborated by BootScan analysis **(Figure 3B)**, which demonstrated a distinct crossover between parental lineages: the recombinant strain showed high similarity to A2 strains sequences across the P1 region and to B3 reference sequences across a part of the P2 region and the entire P3 region.

Visual inspection of the alignment in IGV confirmed uniform read coverage across the genome and no evidence of mixed populations or contamination, excluding co-infection as a potential cause for the observed mosaic signal.

## 4. Discussion

The ongoing evolution of Enterovirus D68, particularly through recombination, represents a critical area of study for understanding its epidemiology, pathogenicity, and potential to cause outbreaks. In this study, we report on the genomic identification and characterization of a novel recombinant EV-D68 strain, A2-Re, detected through genomic surveillance in Maryland in 2025. Our analysis provides compelling evidence that this strain occurred from a recombination event between co-circulating contemporary A2 and historical B3 parental lineages, resulting in a mosaic genome with a P1 region derived from an A2 virus and the P2-P3 regions originating from a B3 virus.

The first indication of this recombinant subclade emerged from a notable phylogenetic inconsistency, while whole-genome analysis placed all 2025 Maryland sequences within the A2 subclade, a distinct monophyletic cluster (A2-Re) displayed shorter branch lengths and formed a separate, well-supported group within A2. These shorter branches indicated reduced genetic divergence, consistent with a recent emergence following a recombination event between A2 and B3 parental lineages, rather than accelerated genetic drift. Such a pattern is characteristic of recombinant viruses, in which different genomic regions follow distinct evolutionary trajectories (37, 38).

Our subgenomic phylogenetic analyses confirmed this pattern, the A2-Re cluster grouped with A2 viruses in the P1 region, but with B3 viruses from Canada and the United States (2022) in the P2–P3 regions. The SimPlot and BootScan analyses supported this topological incongruence, identifying a recombination breakpoint near the 2A/2B junction (∼nt 3,700), a well-known recombination hotspot in enteroviruses (39-41). The recombinant subclade’s limited divergence, together with its tight clustering, suggests a recent origin and localized transmission following the recombination event.

The emergence of A2-Re underscores that recombination remains an active driver of EV-D68 evolution. While previous reports of EV-D68 recombinants have been sporadic (23-26), our detection of a cluster of five nearly identical recombinant genomes in a single season suggests this variant may possess a degree of fitness, allowing for successful transmission and local circulation. At the same time, the detection of only five cases, each identified in different geographic areas, is consistent with silent transmission, low-level community spread that escaped routine clinical detection, likely because infections were mild, asymptomatic, or clinically indistinguishable from other respiratory pathogens. Notably, most previously reported EV-D68 recombination events have occurred in the 5′ untranslated region (5′UTR) (24, 25, 42).

A recent global study on the evolution and transmission dynamics of EV-D68 (27) reported limited recombination events, primarily among Canadian B-genotype, and did not confirm the more frequent recombination patterns described in earlier studies. According to that analysis, the few Canadian sequences showing possible recombination did not exhibit clear phylogenetic signals across the P1, P2, and P3 regions, unlike what was observed in this study.

The functional implications of A2-Re recombination event are substantial. The P1 region encodes the capsid proteins, which are the primary determinants of cell receptor binding, tissue tropism, and antigenicity. Therefore, the A2-derived P1 region suggests that this recombinant may retain the receptor usage and potentially the transmission characteristics of contemporary A2 strains. Conversely, the P2 and P3 regions encode non-structural proteins, including proteases and the RNA polymerase, which are critical for viral replication, immune evasion, and virulence (43, 44). The acquisition of a B3-derived non-structural backbone could potentially alter viral replication kinetics or pathogenicity.

Our findings must be considered in light of certain limitations. The surveillance was conducted within a single health system in Maryland, which may not fully represent the geographic diversity of circulating EV-D68 strains. Furthermore, the identified infections with the recombinant strain were limited, preventing a robust analysis of whether infection with the A2-Re correlates with a distinct clinical phenotype or severity of disease compared to other circulating EV-D68 lineages. A comprehensive analysis of the clinical and epidemiological 2025 EV-D68 outbreak in Maryland, including these recombinant cases, is the subject of an ongoing separate investigation.

## 5. Conclusion

In conclusion, our genomic surveillance in 2025 identified the emergence and local circulation of a novel recombinant EV-D68 strain. This finding has several important implications. First, it demonstrates that recombination between distinct EV-D68 clades is an ongoing evolutionary process that can generate novel, viable Subclade or Clade. Second, it emphasizes the critical importance of whole-genome sequencing and recombination-aware phylogenetic analysis for accurate viral classification. Relying solely on partial genomic sequences (e.g., VP1 only) would have misclassified this cluster as a typical A2 strain. Finally, the continued genetic plasticity of EV-D68 poses a potential challenge for public health, as future recombination events could theoretically generate strains with enhanced transmissibility, neurotropism, or antigenic novelty. Maintained and expanded genomic surveillance is therefore essential to monitor the emergence and spread of such recombinant forms and to understand their clinical impact, particularly their association with severe neurological diseases like AFM.

## Data Availability

All data produced in the present study are available upon reasonable request to the authors

## Funding

This work was supported in part by HHS 75N93021C00045 (HM and AP).

### Disclaimer

*This research was supported [in part] by the Intramural Research Program of the National Institutes of Health (NIH). The contributions of the NIH author(s) are considered Works of the United States Government. The findings and conclusions presented in this paper are those of the author(s) and do not necessarily reflect the views of the NIH or the U*.*S. Department of Health and Human Services*.

